# The Relationship between Continuity of Care with a Primary Care Provider and Duration of Work Disability in Workers with Low Back Pain: A Retrospective Cohort Study

**DOI:** 10.1101/2022.03.06.22271884

**Authors:** Luke R Sheehan, Michael Di Donato, Shannon E Gray, Tyler J Lane, Caryn van Vreden, Alex Collie

**Author notes:** Corresponding author Address: Insurance Work and Health Group, Level 2, 553 St Kilda Road, Melbourne, Victoria, Australia, 3004.

## Abstract

**Objective:** To determine the continuity of care (CoC) provided by general practitioners among workers with low back pain; identify personal, workplace and social factors associated with CoC in this population; and investigate if CoC is associated with working time loss.

**Data sources:** An administrative database containing accepted workers’ compensation claim and service level data, for workers with back pain from five Australian jurisdictions, injured between July 2010 and June 2015.

**Study Design:** A retrospective cohort study. Outcomes were CoC with a general practitioner, measured with the Usual Provider Continuity index, and working time loss, measured as the number of weeks for which workers’ compensation income support benefits were paid.

**Extraction methods:** Eligible workers had at least four general practitioner services, and greater than two weeks working time loss. Usual Provider Continuity index score was categorised as complete, high, moderate, or low CoC. Ordinal logistic regression models examined factors associated with Usual Provider Continuity category. Quantile regression models examined association between duration of working time loss and Usual Provider Continuity category, in four groups with different volumes of general practitioner services.

**Principal Findings:** Complete CoC was observed in 33.8% of workers, high CoC among 37.7%, moderate CoC in 22.1%, and low CoC in 6.4%. Higher Usual Provider Continuity was associated with fewer general practitioner services, older age, living in urban areas, an occupation as a Community and Personal Service Worker or Clerical and Administrative Worker, and the state of Victoria. In workers with more than two months of time loss, those with complete CoC consistently had shorter durations of time loss.

**Conclusions:** Higher CoC with a general practitioner is associated with less working time loss and this relationship is strongest in the sub-acute phase of low back pain.

**Callout Box:** 

**What is known on this topic:**

- Continuity of care is a key component of best practice primary care
- Low back pain is a condition that often requires ongoing management and care from a general practitioner
- The relationship between continuity of care and work disability duration, recovery and return to work in workers with low back pain is not known

**What this study adds:**

- Workers with low back pain who see the same general practitioner for all services (i.e., have a greater continuity of care) generally have shorter durations of working time loss
- Higher continuity of care was observed in workers who had fewer PCP services, were aged over 45, lived in urban areas, and worked as a Community and Personal Service Worker or Clerical and Administrative Worker
- Workers’ compensation systems should consider policies and guidelines that increase continuity of care in injured workers

## Introduction

Continuity of Care (CoC) is an established component of quality primary care ^1–3^. General practitioners (GPs) play a crucial diagnostic and management role with patients experiencing a work-related episode of Low Back Pain (LBP) ^4,5^. This study seeks to explore the relationship between primary care CoC and recovery from an episode of LBP using a novel, multi-state database of Australian workers’ compensation claims and healthcare services data.

GPs play a key role in the diagnosis, treatment, management and care coordination of workers experiencing an episode of LBP ^5^. In Australia, GPs have responsibilities within both the healthcare and workers’ compensation systems. As in the US and Canada, Australian workers’ compensation schemes are cause-based ^6^; a health professional must certify that an injury is work-related before an injured worker can access benefits, and must continue to certify capacity to work for the course of the claim. This primary certification is typically performed by GPs ^7^. GPs are also able to provide access to specialist services that are only available with a GP referral (such as diagnostic imaging). This role as ‘gatekeeper’ to both specialist healthcare services and workers’ compensation benefits means that GPs are not only treatment providers but also have important roles in return to work and healthcare coordination.

Low back pain is a leading cause of disability globally, and occurs at its greatest rates in those of working age ^8^. Work lost to LBP has major economic and personal impacts ^9^. In most cases an episode of LBP resolves within six weeks. For a subset of workers LBP becomes chronic and can lead to long periods of lost working time ^10^. Recovery from LBP is best understood from a perspective that considers biopsychosocial factors alongside injury ‘severity’ as important ^11^. For patients with LBP, highly continuous care with a physical therapist has been linked with lower rates of surgical intervention and lower overall health care costs ^12^ and continuity with a health care provider is also desired by patients with LBP ^13^. Recovery and return to work for an episode of LBP is associated with a range of occupational, jurisdictional, social and demographic factors ^14,15^. CoC with a GP is hypothesised to be related to recovery and return to work, but this relationship remains unexplored. This study combines administrative claim data with administrative healthcare payments data, and applies a CoC metric to provide new insight into the relationship between patient-physician continuity and recovery from LBP.

This study has three primary research questions:

1. How continuous is the care provided by GPs to workers with accepted workers’ compensation claims for LBP?
2. What demographic, occupational and social factors are associated with GP CoC in workers with LBP in workers’ compensation systems?
3. What is the association between GP CoC and the duration of working time lost among workers with LBP in workers’ compensation systems?

## Methods

### Setting

There are eleven workers’ compensation jurisdictions in Australia: one for each of the six states and two territories, and three for national industries and employers ^16^. Australian workers’ compensation schemes provide wage replacement payments and ‘reasonable and necessary’ medical expenses and services for workers who have suffered an injury or illness in the course of their employment ^16^. Workers need to provide medical certificates from their doctors demonstrating work capacity impairments in order to continue receiving income support payments. In Australian workers’ compensation systems, GPs have both an administrative role (medical certification) as well as a treatment role (supporting recovery and return to work). There are no restrictions around access to particular GPs; the choice of provider is at the patient’s discretion.

### Data Source

This study used data from the Monash University Multi-Jurisdictional Workers’ Compensation Database, which has been previously described ^17^. The database contains de-identified administrative workers’ compensation claims for musculoskeletal conditions and associated service payments from five of Australia’s workers’ compensation jurisdictions, with joint coverage of approximately 60% of the national labour force. The database contains claims with date of injury ranging from 1 July 2010 to 30 June 2015.

### Sample/ Inclusion Criteria

Included claims were for LBP, with a minimum of four recorded GP service payments, greater than two weeks paid time loss, and from the workers’ compensation schemes for the states of Queensland, Victoria, and South Australia. The two other jurisdictions in the data base were excluded as their services data did not contain a unique GP identifier enabling calculation of CoC metrics.

The three included jurisdictions comprised 52% of the Australian labour force in 2013 (the mid-point of the study) ^18^. Claims with less than four GP services were excluded, consistent with other studies of CoC ^19–21^. LBP claims were defined using Nature of Injury and Location of Injury codes from the Type of Occurrence Classification System version 3.1 (Supplementary Table 1) ^22^. Only claims with over two weeks’ time loss were included as in some jurisdictions, claims with less than two weeks’ time loss are managed and paid for by the employer and not the workers’ compensation authority ^16^.

### General Practitioner Services

GP services were defined as payments from the workers’ compensation scheme to a GP for services that included a patient interaction. Other payments such as for report writing, review of reports or programs, provision of medication, or for patient non-attendance were excluded. The database contains a unique and de-identified code for each GP, enabling services provided by the same GP to be quantified.

### Continuity of Care

Continuity of care with a GP was measured using the Usual Provider Continuity (UPC) index^23^. The UPC is calculated as the proportion of GP services that were with the most frequently seen service provider and has a range from 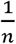 (all services with different GP) to 1 (all services with the same GP). The UPC index was chosen as it is the most direct measure of the relationship between worker and their ‘usual’ GP, and is not affected by the dispersion of other GPs a worker may have visited. UPC has been shown to have non-linear associations with various outcomes and so was classified into previously used categories ^21^ for analysis: complete CoC (UPC = 1), high CoC (UPC score of 0.75-0.99), moderate CoC (UPC score of 0.5-0.74), and low CoC (UPC <0.5).

### Working Time Loss

Working time loss was defined as the number of weeks of income support payments paid. There were differences in how jurisdictions recorded a weekly payment (either 5 or 7 days), these were standardised across jurisdictions to 7 days. The number of days paid for each claim was then summed and divided by 7. Time loss is therefore measured in paid calendar weeks. Time loss duration was right censored at 104 weeks, which is when workers in some jurisdictions begin to lose access to benefits.

### Covariates

GP service count was categorised into quartiles for analysis with four service groups defined: low service group (4-6 GP services), moderate service group (7-11 GP services), high service group (12-22 GP services), and very high service group (23+ GP services). Jurisdiction refers to the state in which the worker made their workers’ compensation claim. Age was categorised into 10-year groups. Sex was provided as either male or female. Occupation was available as Major Groups as defined by the Australian and New Zealand Standard Classification of Occupations ^24^. Remoteness was defined by matching postcode information to the Australian Statistical Geography Standard, with Remote and Very Remote combined into a single group due to low frequency in these categories ^25^. There were 836 cases with missing data for Remoteness (4.6% of the sample). Multiple imputation for missing remoteness was performed using ordinal logistic regression.

### Analysis

Factors associated with UPC score were estimated using generalised ordered logistic regression with partial parallel odds ^26^. This regression model is a variation of ordered logistic regression in which covariates that have parallel odds across levels are estimated once (as in standard ordered regression), while covariates that do not have parallel odds across comparisons have unique estimates at each level (similar to multinomial regression). Covariates (as defined above) were factors that were hypothesised to have a relationship with UPC values, and were available in the harmonised administrative data.

Analysis of the relationship between CoC and duration of working time loss was structured to take into account that workers with less time loss were more likely to have higher UPC scores simply because they were also more likely to have had fewer GP services. To minimise this confounding, analysis was performed separately for each GP service count quartile.

Kaplan-Meier failure plots (that present the cumulative proportion of workers by their time loss duration) were used to assess the relationship between CoC and working time loss. The Kaplan-Meier plots (Figure 1) indicated that the relationship between UPC category and time loss was not constant over time and for this reason, Cox regression was not considered appropriate (due to non-proportional hazards). Quantile regression models were developed in each GP service quartile to assess the relationship between CoC and time loss. Quantile regression is similar to least squares estimation for the mean in linear regression models, but adapted to estimate quantiles instead of the mean ^27^. As estimates are calculated separately at each quantile, the relationship between covariates and the outcome are no longer constrained to be constant over time, as they are in Cox regression.

**Figure 1.**
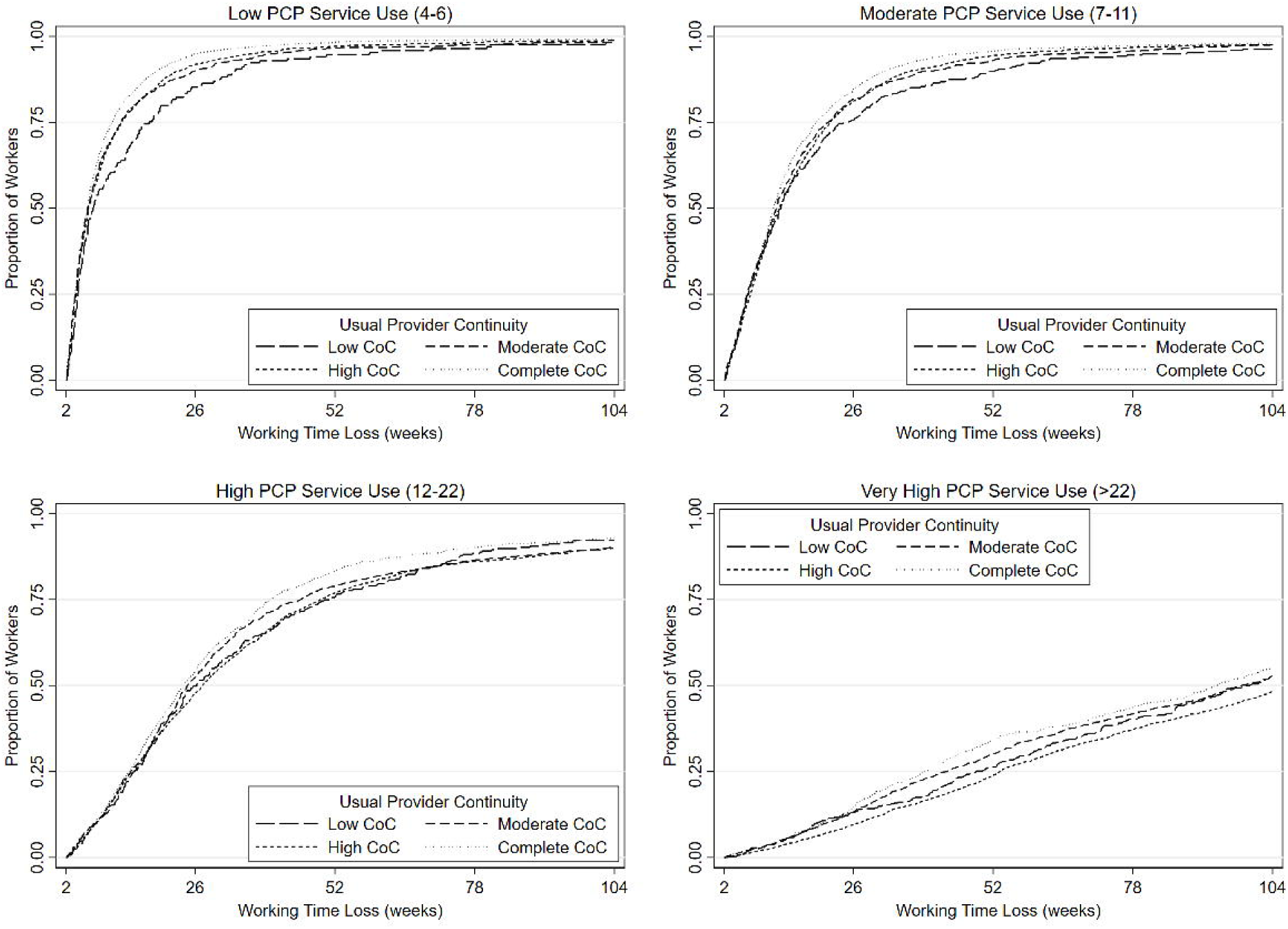
Working Time Loss Duration by Usual Provider Category (UPC) and General Practitioner (GP) Service Use Quartile. Kaplan-Meier failure function for time spent on weekly benefits - graphed by GP service use quartile (between plots) and UPC category (within plots).

The outcomes for the quantile regression models were weeks of working time loss at q10, q25, q50, q75, and q90. The independent variable of interest was the UPC category. Covariates included GP service count as a continuous variable, jurisdiction, age, sex, occupation, and remoteness (as previously defined).

All statistical analysis was performed using Stata 16 ^28^, with the user-written program gologit2 used for generalised ordered logit regression ^26^.

### Sensitivity Analysis

A sensitivity analysis using the Bice-Boxerman Continuity of Care Index (COCI) in place of the UPC as the measure of CoC was conducted. The COCI metric measures the degree dispersal of services among different providers,^29^ unlike the UPC which only measures the continuity with the most common provider. The COCI was categorised in the same way as the UPC (as has been used previously).^21^ Analysis performed was identical to that for the UPC metric.

## Results

### Sample Characteristics

There were 18,696 claims that met the eligibility criteria, with an associated 305,976 GP services. There were 13,276 GPs who provided at least one service to a worker in the sample. The median number of services provided by a GP was 9 with an IQR of 3-27 and a maximum of 1,240. Workers from Queensland and Victoria comprised the majority of the sample with 42.7% and 38.9% of claims respectively, the remainder were from South Australia (18.4%) [Table 1]. Included workers had a median of 11 (IQR: 6-22) GP services. The median duration of working time loss was 18.0 weeks (IQR: 7.1-52.0). Over half the sample were between 35 and 55 years old, 63.0% were male, and two thirds of claims were from the major cities of Australia.

**Table 1.** Sample characteristics

| Variable | Complete Continuity of Care<br>UPC 1<br>(column %) | High Continuity of Care<br>UPC 0.75-0.99<br>(column %) | Moderate Continuity of Care<br>UPC 0.5-0.75<br>(column %) | Low Continuity of Care<br>UPC <0.5<br>(column %) | Total<br>(column %) |
| --- | --- | --- | --- | --- | --- |
| **UPC category (row %)** | 6 389 (34.2%) | 7 004 (37.5%) | 4 114 (22.0%) | 1 189 (6.4%) | 18 696 (100.0%) |
| **Number of general practitioner services (quartiles)** |  |  |  |  |  |
| 4-6 (Low service group) | 2 652 (41.5%) | 1 115 (15.9%) | 867 (21.1%) | 169 (14.2%) | 4 803 (25.7%) |
| 7-11 (Moderate service group) | 1 942 (30.4%) | 1 489 (21.3%) | 1 090 (26.5%) | 295 (24.8%) | 4 816 (25.8%) |
| 12-22 (High service group) | 1 162 (18.2%) | 1 981 (28.3%) | 1 062 (25.8%) | 295 (24.8%) | 4 500 (24.1%) |
| 23+ (Very high service group) | 633 (9.9%) | 2 419 (34.5%) | 1 095 (26.6%) | 430 (36.2%) | 4 577 (24.5%) |
| **Age** |  |  |  |  |  |
| 15-25 years | 698 (10.9%) | 658 (9.4%) | 505 (12.3%) | 165 (13.9%) | 2 026 (10.8%) |
| 26-35 years | 1 278 (20.0%) | 1 516 (21.6%) | 1 094 (26.6%) | 354 (29.8%) | 4 242 (22.7%) |
| 36-45 years | 1 671 (26.2%) | 1 952 (27.9%) | 1 115 (27.1%) | 319 (26.8%) | 5 057 (27.0%) |
| 46-55 years | 1 733 (27.1%) | 1 905 (27.2%) | 952 (23.1%) | 254 (21.4%) | 4 844 (25.9%) |
| 56-65 years | 941 (14.7%) | 926 (13.2%) | 423 (10.3%) | 88 (7.4%) | 2 378 (12.7%) |
| 66+ years | 68 (1.1%) | 47 (0.7%) | 25 (0.6%) | 9 (0.8%) | 149 (0.8%) |
| **Sex** |  |  |  |  |  |
| Male | 3 910 (61.2%) | 4 501 (64.3%) | 2 586 (62.9%) | 780 (65.6%) | 11 777 (63.0%) |
| Female | 2 479 (38.8%) | 2 503 (35.7%) | 1 528 (37.1%) | 409 (34.4%) | 6 919 (37.0%) |
| **Jurisdiction** |  |  |  |  |  |
| Queensland | 3 136 (49.1%) | 2 597 (37.1%) | 1 772 (43.1%) | 487 (41.0%) | 7 992 (42.7%) |
| Victoria | 2 308 (36.1%) | 3 192 (45.6%) | 1 407 (34.2%) | 366 (30.8%) | 7 273 (38.9%) |
| South Australia | 945 (14.8%) | 1 215 (17.3%) | 935 (22.7%) | 336 (28.3%) | 3 431 (18.4%) |
| **Remoteness** |  |  |  |  |  |
| Major Cities of Australia | 4 465 (69.9%) | 4 731 (67.5%) | 2 628 (63.9%) | 669 (56.3%) | 12 493 (66.8%) |
| Inner Regional Australia | 1 130 (17.7%) | 1 352 (19.3%) | 826 (20.1%) | 256 (21.5%) | 3 564 (19.1%) |
| Outer Regional Australia | 520 (8.1%) | 551 (7.9%) | 371 (9.0%) | 154 (13.0%) | 1 596 (8.5%) |
| Remote/Very Remote Australia | 25 (0.4%) | 44 (0.6%) | 36 (0.9%) | 20 (1.7%) | 125 (0.7%) |
| Missing | 249 (3.9%) | 326 (4.7%) | 253 (6.1%) | 90 (7.6%) | 918 (4.9%) |
| **Occupation** |  |  |  |  |  |
| Labourers | 1 714 (26.8%) | 1 899 (27.1%) | 1 219 (29.6%) | 335 (28.2%) | 5 167 (27.6%) |
| Community and Personal Service Workers | 1 350 (21.1%) | 1 264 (18.0%) | 751 (18.3%) | 219 (18.4%) | 3 584 (19.2%) |
| Machinery Operators and Drivers | 1 039 (16.3%) | 1 267 (18.1%) | 657 (16.0%) | 221 (18.6%) | 3 184 (17.0%) |
| Technicians and Trades Workers | 1 050 (16.4%) | 1 192 (17.0%) | 703 (17.1%) | 208 (17.5%) | 3 153 (16.9%) |
| Professionals | 558 (8.7%) | 549 (7.8%) | 344 (8.4%) | 79 (6.6%) | 1 530 (8.2%) |
| Managers | 267 (4.2%) | 316 (4.5%) | 170 (4.1%) | 56 (4.7%) | 809 (4.3%) |
| Sales Workers | 220 (3.4%) | 295 (4.2%) | 154 (3.7%) | 49 (4.1%) | 718 (3.8%) |
| Clerical and Administrative<br>Workers | 191 (3.0%) | 222 (3.2%) | 116 (2.8%) | 22 (1.9%) | 551 (2.9%) |

UPC: Usual Provider Continuity

Complete CoC was observed in 6,389 (34.2%) of workers, high CoC in 7,004 (37.5%), moderate CoC in 4,114 (22.0%), and low CoC in 1,189 (6.4%) [Table 1].

### Factors Associated with Continuity of Care

The number of GP services was strongly associated with UPC category with more GP services associated with lower UPC (Table 2). The largest effects were seen for complete continuity: in comparison to workers with 4-6 GP services, those with 7-11 services had 47% lower odds of complete continuity (OR: 0.53, 95% CI: 0.49-0.57), those with 12-22 GP services had 74% lower odds (OR: 0.26, 95% CI: 0.24-0.29), and those with over 23 GP services had 89% lower odds (OR: 0.11, 95% CI: 0.10-0.13).

**Table 2.**
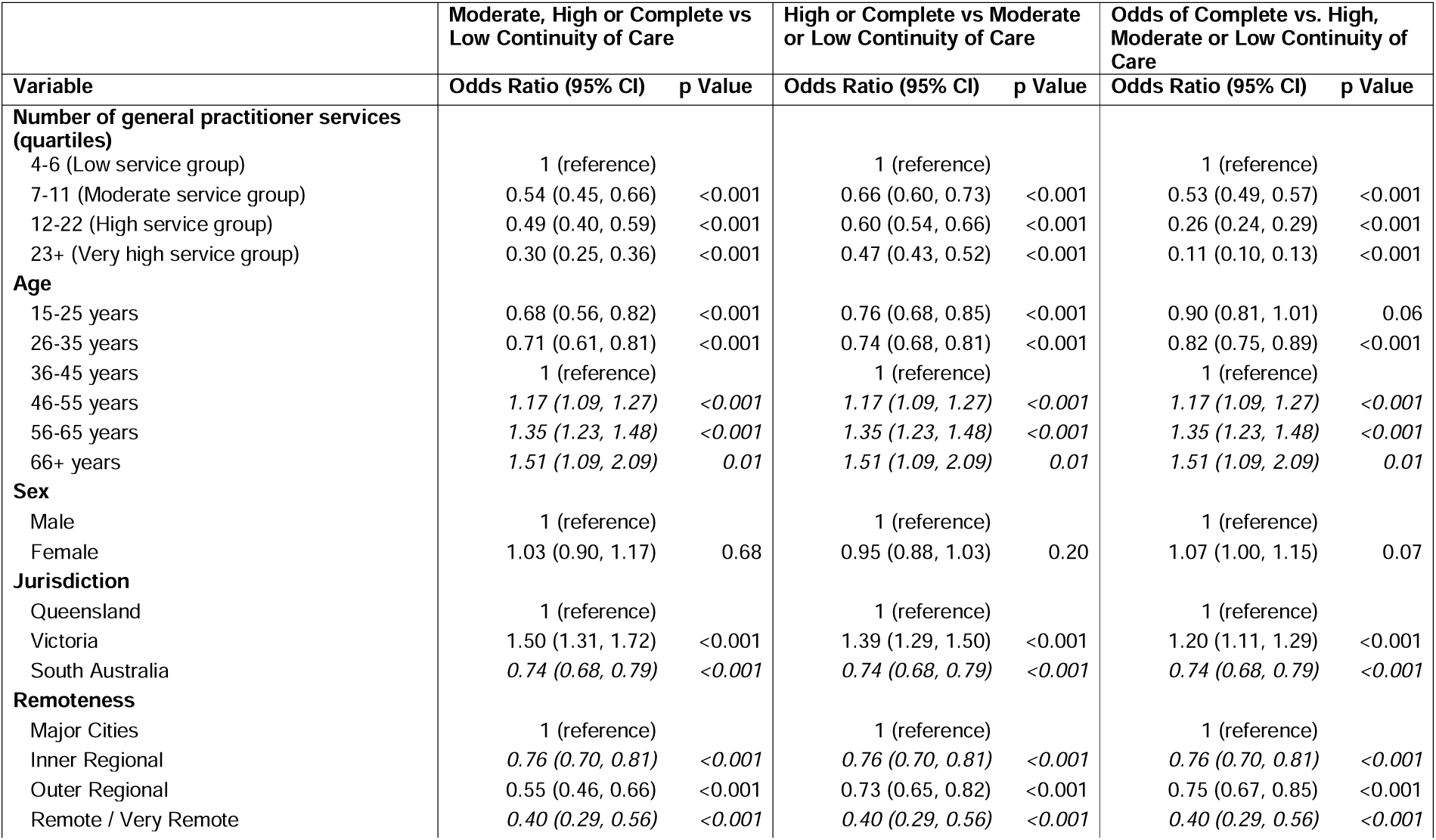
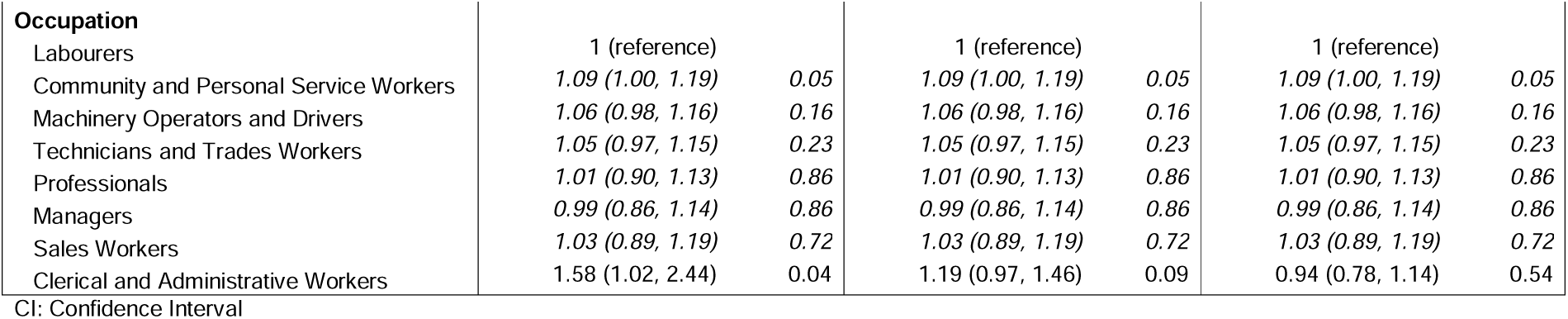
Generalised ordered logit partial proportional odds models with Usual Provider Continuity category as the outcome. Variables with estimates in italics have a single estimate applied across each combination. Variables in plain text have estimates for each group.

Age had a strong positive relationship with UPC score, with odds ratios increasing in every age bracket after 25 years (Table 2). Workers in the state of Victoria had higher odds and those from the state of South Australia had lower odds of higher UPC scores, when compared to workers from the state of Queensland. The odds of higher UPC scores was largest in the major cities and decreased with remoteness.

Occupation generally did not have a strong relationship with UPC score (Table 2), but Community and Personal Service Workers did have better odds of higher UPC score when compared to Labourers. Clerical and Administrative Workers had significantly better odds than Labourers of not having a low UPC score, but did not differ at other levels.

### Relationship between Continuity of Care and Working Time Loss

Figure 1 plots the cumulative proportion of weeks of working time loss (Kaplan-Meier failure function) by each GP quartile and UPC category. There are substantial differences in working time loss between the GP service groups, with the duration of weekly benefits strongly related to the volume of GP service use. There is a lack of variation between the UPC categories for shorter durations of time loss, before they begin to diverge.

#### Low GP service group

Quantile regression found that among workers recording 4 to 6 GP services, at the 75^th^ and 90^th^ percentiles all UPC categories had significantly longer time loss duration than those with complete continuity, and there was a consistent pattern of a lower UPC score corresponding to a longer duration of time loss (Table 3).

**Table 3.** Difference in time loss for low, moderate and high continuity groups, compared to claims with complete continuity estimated from quantile regression (models were also adjusted for jurisdiction, age, sex, remoteness, and occupation)

| Percentile | Complete continuity of care<br>UPC = 1 | High continuity of care<br>UPC 0.75 – 0.99 |  | Moderate continuity of care<br>UPC 0.5 – 0.74 |  | Low continuity of care<br>UPC <0.5 |  |
| --- | --- | --- | --- | --- | --- | --- | --- |
|  | Weeks<sup>#</sup> | Coefficient<sup>^</sup> (95% CI) | p Value | Coefficient<sup>^</sup> (95% CI) | p Value | Coefficient<sup>^</sup> (95% CI) | p Value |
| **Low GP Service Group (4-6 GP services)** |  |  |  |  |  |  |  |
| 10<sup>th</sup> | 2.40 | -0.04 (-0.20, 0.12) | 0.65 | 0.00 (-0.12, 0.12) | 1.00 | 0.17 (-0.30, 0.64) | 0.48 |
| 25<sup>th</sup> | 2.95 | 0.01 (-0.19, 0.20) | 0.95 | -0.22 (-0.43, -0.02) | 0.03 | 0.11 (-0.41, 0.62) | 0.62 |
| 50<sup>th</sup> | 4.30 | 0.11 (-0.21, 0.42) | 0.51 | 0.02 (-0.40, 0.44) | 0.91 | 0.80 (-0.76, 2.37) | 0.31 |
| 75<sup>th</sup> | 7.90 | 0.97 (0.26, 1.68) | 0.01 | 1.12 (0.09, 2.14) | 0.03 | 5.98 (3.49, 8.47) | <0.001 |
| 90<sup>th</sup> | 15.75 | 3.23 (0.92, 5.54) | 0.01 | 7.06 (3.09, 11.02) | <0.001 | 14.93 (8.49, 21.36) | <0.001 |
| **Moderate GP Service Group (7-11 GP services)** |  |  |  |  |  |  |  |
| 10<sup>th</sup> | 2.71 | -0.12 (-0.50, 0.26) | 0.54 | -0.19 (-0.59, 0.22) | 0.36 | 0.73 (0.08, 1.38) | 0.03 |
| 25<sup>th</sup> | 4.05 | 0.16 (-0.38, 0.70) | 0.56 | 0.12 (-0.44, 0.67) | 0.68 | 0.29 (-0.54, 1.13) | 0.49 |
| 50<sup>th</sup> | 6.71 | 1.08 (0.26, 1.90) | 0.01 | 0.86 (0.19, 1.54) | 0.01 | 0.77 (-0.90, 2.45) | 0.37 |
| 75<sup>th</sup> | 11.46 | 1.46 (0.05, 2.87) | 0.04 | 2.54 (0.95, 4.13) | 0.002 | 6.64 (2.70, 10.59) | 0.001 |
| 90<sup>th</sup> | 19.46 | 3.52 (0.07, 6.97) | 0.05 | 6.02 (2.04, 10.00) | 0.003 | 26.05 (13.05, 39.05) | <0.001 |
| **High GP Service Group (12-22 GP services)** |  |  |  |  |  |  |  |
| 10<sup>th</sup> | 5.34 | 0.56 (-0.66, 1.79) | 0.37 | -0.33 (-1.47, 0.80) | 0.57 | 0.59 (-1.55, 2.73) | 0.59 |
| 25<sup>th</sup> | 8.30 | 0.44 (-0.88, 1.78) | 0.51 | 0.08 (-1.50, 1.67) | 0.92 | 1.76 (-0.67, 4.19) | 0.16 |
| 50<sup>th</sup> | 13.21 | 1.84 (0.31, 3.37) | 0.02 | 0.33 (-1.54, 2.20) | 0.73 | 4.11 (0.99, 7.23) | 0.01 |
| 75<sup>th</sup> | 20.76 | 2.48 (-0.76, 5.71) | 0.13 | 1.20 (-2.67, 5.08) | 0.54 | 7.12 (1.20, 13.04) | 0.02 |
| 90<sup>th</sup> | 42.48 | 5.70 (-0.18, 11.59) | 0.06 | 3.45 (-2.51, 9.41) | 0.26 | 3.00 (-6.75, 12.74) | 0.55 |
| **Very High GP Service Group (23+ GP services)** |  |  |  |  |  |  |  |
| 10<sup>th</sup> | 10.66 | 6.24 (2.12, 10.37) | 0.003 | 0.98 (-3.84, 5.80) | 0.69 | -2.64 (-9.75, 4.48) | 0.47 |
| 25<sup>th</sup> | 16.31 | 12.51 (6.66, 18.36) | <0.001 | 3.67 (-2.61, 9.95) | 0.25 | 8.16 (-0.05, 16.38) | 0.05 |
| 50<sup>th</sup> | 44.57 | 8.15 (3.80, 12.49) | <0.001 | 3.38 (-2.92, 9.67) | 0.29 | 1.32 (-6.62, 9.26) | 0.75 |

Note: # Weeks lost at this percentile calculated at the reference value of categorical variables and minimum number of GP services.
Note: ^ Coefficient refers to difference in weeks lost compared to those with complete continuity of care (UPC = 1)
UPC: Usual Provider Continuity, CI: Confidence interval, GP: General practitioner

#### Moderate GP service group

Among workers recording between 7 and 11 GP services, quantile regression identified that workers with moderate and high UPC scores had an additional week in duration of time loss at the median time loss (compared to complete continuity) [Table 3]. At the 75^th^ and 90th percentiles all UPC categories had significantly longer duration of time loss than workers with complete continuity, and a consistent pattern of a lower UPC score corresponding to a longer time loss.

#### High GP service group

Quantile regression among workers with 12 to 22 recorded GP services found there were significantly longer durations of time loss among workers with low and high UPC scores at the 50^th^ percentile. Workers with low UPC scores also recorded significantly longer durations at the 75^th^ percentile with an increase of 7.12 weeks (95% CI: 1.20-13.04).

#### Very high GP service group

Among workers with 23 or more GP services, those with a high UPC score had significantly longer durations of time loss at all percentiles (Table 3). Analysis of the 75^th^ and 90^th^ percentiles were not performed as time loss at these percentiles went beyond 104 weeks.

### Sensitivity Analysis

Detailed results of the sensitivity analysis using the COCI instead of the UPC are reported in the Supplementary Materials. Factors associated with CoC were similar regardless of whether UPC or COCI was used to measure CoC. The relationship between CoC and working time loss showed similar trends for COCI as with UPC, although the effect sizes were generally smaller.

## Discussion

This study investigates the relationship between continuity of care (CoC) with a general practitioner (GP) and the duration of working time loss in Australian workers with accepted workers’ compensation claims for low back pain (LBP). We observed a largely consistent direction of effect in that workers who consulted the same GP in all services during their compensation claim (complete CoC) had shorter durations of working time loss while workers with low CoC generally had the longest durations of working time loss. The results provide evidence that more continuous care is associated with shorter durations of time off work and may facilitate return to work in workers with low back pain claims.

The study results demonstrate that CoC begins to influence working time loss in workers whose claims exceed one to two months duration. This is consistent with the natural history of LBP in which most acute and sub-acute cases resolve within six to eight weeks ^30^. After this acute period, pain becomes less predictive of time loss ^14^ and psychosocial factors are more strongly associated with outcomes including return to work. Our findings suggest that as LBP progresses to a chronic phase, an ongoing relationship with a single GP becomes a more important factor in worker recovery and return to work.

The risk of receiving different or conflicting treatment and advice when consulting multiple GPs is a potential explanation for this association. Despite well-documented guidelines, treatment of LBP by GPs does not always adhere to best practice ^31–33^. There is some evidence for this relationship in physical therapy, where lower CoC with physical therapists was associated with negative outcomes including higher incidence of invasive surgery and higher healthcare costs ^12^.

Another potential explanation for the observed association between CoC and duration of working time loss is that workers consulting multiple GPs will be asked to describe and quantify their pain experience repeatedly. As LBP is a symptomatic condition often with no externally observable features, a worker will need to describe their pain and its relationship to the workplace in order for a GP to provide certification and recommend treatment. The consequences of both having to ‘justify’ their need to be off work and relive the experience(s) that led to injury can be counterproductive to the psychosocial measures that promote recovery from an episode of LBP ^9^, and may contribute to workers adopting a ‘sick-role’ that can contribute to delayed recovery ^34^.

The relationship between CoC and working time loss observed in this study may not be entirely explained by the impact of CoC. Patients who are not recovering from their injury may become dissatisfied and more likely to seek out different GPs. A lack of satisfaction with their most recent GP visit was a strong predictor of not having a ‘regular’ GP in one Australian study ^35^.

The present study also identified worker characteristics that could be utilised to target workers who are at greater risk of experiencing lower CoC in this cohort. Higher continuity was observed in workers from major cities than in workers from regional areas. Availability and retention of GPs in regional areas is known to be limited in Australia ^36^. Additionally there are fewer allied health professionals per person in regional Australia ^37^ meaning patient dependence on GPs for the treatment of LBP is likely to be greater.

There was a consistent pattern of higher CoC with increasing age. Older workers are more likely to have a regular GP ^35^, and are likely to see that GP from the beginning of management of their work-related LBP. Younger workers are less likely to already have a regular GP, so they may visit any GP who is available or visit multiple GPs before they find a GP they feel comfortable with.

The state in which a worker filed their workers’ compensation claim had a significant impact on CoC, suggesting that state differences in policy may be one factor affecting the volume of GP visits seen in this cohort. In South Australia and Queensland, certificates of capacity must be completed by a medical practitioner ^38,39^. In Victoria the first certificate of capacity must be completed by a medical practitioner, but subsequent certificates can be completed by other allied health professionals, such as a physiotherapist or chiropractor ^40^. While it is reasonable for a jurisdiction to require a worker to be certified as unable to work, if the worker’s usual GP is not available another GP may be consulted. It is unlikely that all GPs would simply perform an administrative service when consulted by an injured worker ^41^, and this introduces the potential for the worker to receive conflicting advice, alterations to treatment, and increases the likelihood of having to relive the injury experience. Policies that enable or encourage workers to see the same GP, or limit the need for workers to consult multiple GPs, may ultimately be beneficial for workers’ compensation systems by reducing working time loss and associated costs.

The strengths of this study include use of a large, novel database from multiple Australian workers’ compensation jurisdictions. The combination of claim-level and service-level data enabled analysis of outcomes that have not previously been researched in Australian workers’ compensation systems. A sensitivity analysis with another measure of CoC indicates that results were robust. The use of workers’ compensation data enables GP CoC to be examined for a single condition, as only services funded by workers’ compensation are available in the data set. However, this also means that any GP services paid for by the worker are not captured in the data set and unable to be included in the CoC calculation.

This is important to note in Australia where healthcare is relatively accessible. The database did not enable identification of weekly payments for partial return to work, so working time loss may be overestimated. Covariates were limited to those available in the administrative data sets, so clinical information such as pain severity was not able to be incorporated in analyses. There are also limitations to the ability of the UPC metric to capture CoC. UPC was calculated retrospectively and analysed as a constant value. In reality, CoC (and the UPC metric) is something that develops and changes over time. The UPC metric also does not capture how the patient actually experiences CoC, the experience of seeing multiple GPs may depend on information flow between different GPs, or whether consultation with different GPs is a choice or necessity.

## Conclusion

Workers with accepted workers’ compensation claims for LBP spent less time off work when they had higher CoC. This effect was most prominent after one to two months of working time loss and persisted after adjustment for age, sex, jurisdiction, remoteness, and occupation. This new evidence suggests that CoC with a GP has greatest impact during the sub-acute phase of LBP and may play a role in preventing LBP from becoming persistent. Workers’ compensation systems can introduce policies and programs that facilitate greater continuity of care and educate and encourage workers about the importance of CoC with a GP as part of recovery and return to work. While this paper is focused on a specific cohort, given the large global burden of disability attributable to LBP, the findings warrant investigation of primary care CoC in broader settings.

## Data Availability

Data used in this paper was provided to the authors by the workers' compensation authorities and are not available for distribution by the authors.

## Acknowledgments

The following organisations supplied data for use in this study: WorkCover Queensland, WorkSafe Victoria, and ReturnToWork South Australia. The views expressed in this paper are those of the authors and are not necessarily the views of the data providers or study funders. Data used in this paper was provided to the authors by the workers’ compensation authorities and are not available for distribution by the authors. The authors would like to acknowledge the assistance of Ms Dianne Beck in the cleaning and preparation of data for this paper.

Funding for this project was provided by a Discovery Project Grant (DP190102473) from the Australian Research Council and a grant from Safe Work Australia.

