## Supplementary Tables for "The Relationship between Continuity of Care with a Primary Care Provider and Duration of Work Disability in Workers with Low Back Pain: A Retrospective Cohort Study"

**Supplementary Table 1.** Type of occurrence classification system version 3.1 codes used to define Low Back Pain

| Nature of Injury | 228 – Trauma to muscles and tendons, not elsewhere classified  229 – Trauma to muscles and tendons, unspecified  239 – Soft tissue injuries due to trauma or unknown mechanisms with insufficient information to code elsewhere  422 – Disc displacement, prolapse, degeneration, or hernia  459 – Back pain, lumbago, and sciatica  533 – Muscle / tendon strain (non-traumatic) |
| --- | --- |
| Location of Injury | 311 – Lower back |
| Mechanism of Injury | Any |
| Agency of Injury | Any |

**Supplementary Table 2.** Generalised ordered logit partial proportional odds models with Bice-Boxerman Continuity of Care Index category as the outcome. Variables with estimates in italics have a single estimate applied across each combination. Variables in plain text have estimates for each group.

|  | **Moderate, High or Complete vs Low Continuity of Care** | | **High or Complete vs Moderate or Low Continuity of Care** | | **Odds of Complete vs. High, Moderate or Low Continuity of Care** | |
| --- | --- | --- | --- | --- | --- | --- |
| **Variable** | **Odds Ratio (95% CI)** | **p Value** | **Odds Ratio (95% CI)** | **p Value** | **Odds Ratio (95% CI)** | **p Value** |
| **Number of General Practitioner services (quartiles)** |  |  |  |  |  |  |
| 4-6 (Low service group) | 1 (reference) |  | 1 (reference) |  | 1 (reference) |  |
| 7-11 (Moderate service group) | 0.67 (0.58, 0.78) | <0.001 | 0.76 (0.63, 0.93) | 0.01 | 0.69 (0.62, 0.76) | <0.001 |
| 12-22 (High service group) | 0.59 (0.31, 1.14) | 0.09 | 0.56 (0.38, 0.81) | 0.01 | 0.41 (0.17, 0.97) | 0.05 |
| 23+ (Very high service group) | 0.56 (0.41, 0.76) | 0.004 | 0.52 (0.46, 0.59) | <0.001 | 0.14 (0.11, 0.18) | <0.001 |
| **Age** |  |  |  |  |  |  |
| 15-25 years | 0.75 (0.67, 0.84) | <0.001 | 0.80 (0.72, 0.89) | <0.001 | 0.90 (0.80, 1.01) | 0.06 |
| 26-35 years | 0.77 (0.65, 0.84) | <0.001 | 0.78 (0.72, 0.84) | <0.001 | 0.79 (0.72, 0.88) | <0.001 |
| 36-45 years | 1 (reference) |  | 1 (reference) |  | 1 (reference) |  |
| 46-55 years | *1.15 (1.07, 1.24)* | *<0.001* | *1.15 (1.07, 1.24)* | *<0.001* | *1.15 (1.07, 1.24)* | *<0.001* |
| 56-65 years | *1.33 (1.21, 1.45)* | *<0.001* | *1.33 (1.21, 1.45)* | *<0.001* | *1.33 (1.21, 1.45)* | *<0.001* |
| 66+ years | *1.59 (1.16, 2.17)* | *0.004* | *1.59 (1.16, 2.17)* | *0.004* | *1.59 (1.16, 2.17)* | *0.004* |
| **Sex** |  |  |  |  |  |  |
| Male | 1 (reference) |  | 1 (reference) |  | 1 (reference) |  |
| Female | 1.00 (0.92, 1.08) | 0.93 | 1.08 (1.00, 1.15) | 0.04 | 1.05 (0.97, 1.12) | 0.21 |
| **Jurisdiction** |  |  |  |  |  |  |
| Queensland | 1 (reference) |  | 1 (reference) |  | 1 (reference) |  |
| Victoria | 1.41 (1.28, 1.55) | <0.001 | 1.33 (1.22, 1.45) | <0.001 | 1.14 (1.00, 1.31) | 0.05 |
| South Australia | *0.74 (0.68, 0.80)* | *<0.001* | *0.74 (0.68, 0.80)* | *<0.001* | *0.74 (0.68, 0.80)* | *<0.001* |
| **Remoteness** |  |  |  |  |  |  |
| Major Cities | 1 (reference) |  | 1 (reference) |  | 1 (reference) |  |
| Inner Regional | *0.75 (0.70, 0.80)* | *<0.001* | *0.75 (0.70, 0.80)* | *<0.001* | *0.75 (0.70, 0.80)* | *<0.001* |
| Outer Regional | *0.74 (0.66, 0.81)* | *<0.001* | *0.74 (0.66, 0.81)* | *<0.001* | *0.74 (0.66, 0.81)* | *<0.001* |
| Remote / Very Remote | *0.41 (0.30, 0.57)* | *<0.001* | *0.41 (0.30, 0.57)* | *<0.001* | *0.41 (0.30, 0.57)* | *<0.001* |
| **Occupation** |  |  |  |  |  |  |
| Labourers | 1 (reference) |  | 1 (reference) |  | 1 (reference) |  |
| Community and Personal Service Workers | *1.09 (1.00, 1.18)* | *0.04* | *1.09 (1.00, 1.18)* | *0.04* | *1.09 (1.00, 1.18)* | *0.04* |
| Machinery Operators and Drivers | *1.08 (1.00, 1.17)* | *0.06* | *1.08 (1.00, 1.17)* | *0.06* | *1.08 (1.00, 1.17)* | *0.06* |
| Technicians and Trades Workers | *1.09 (1.00, 1.18)* | *0.05* | *1.09 (1.00, 1.18)* | *0.05* | *1.09 (1.00, 1.18)* | *0.05* |
| Professionals | *1.02 (0.91, 1.14)* | *0.73* | *1.02 (0.91, 1.14)* | *0.73* | *1.02 (0.91, 1.14)* | *0.73* |
| Managers | *1.02 (0.89, 1.18)* | *0.73* | *1.02 (0.89, 1.18)* | *0.73* | *1.02 (0.89, 1.18)* | *0.73* |
| Sales Workers | *1.02 (0.88, 1.17)* | *0.83* | *1.02 (0.88, 1.17)* | *0.83* | *1.02 (0.88, 1.17)* | *0.83* |
| Clerical and Administrative Workers | *1.04 (0.88, 1.22)* | *0.66* | *1.04 (0.88, 1.22)* | *0.66* | *1.04 (0.88, 1.22)* | *0.66* |

CI: Confidence Interval

**Supplementary Table 3.** Difference in time loss for low, moderate and high continuity groups, compared to claims with complete continuity estimated from quantile regression (models were also adjusted for jurisdiction, age, sex, remoteness, and occupation)

| **Percentile** | **Complete continuity of care**  **COCI = 1** | **High continuity of care**  **COCI 0.75 – 0.99** | | **Moderate continuity of care**  **COCI 0.5 – 0.74** | | **Low continuity of care**  **COCI <0.5** | |
| --- | --- | --- | --- | --- | --- | --- | --- |
|  | Weeks^#^ | Coefficient^ (95% CI) | p Value | Coefficient^ (95% CI) | p Value | Coefficient^ (95% CI) | p Value |
| **Low GP Service Group (4-6 GP services)** | | | | | | | |
| 10^th^ | 2.40 | N/A^*^ | | -0.03 (-0.19, 0.13) | 0.71 | 0.03 (-0.10, 0.17) | 0.62 |
| 25^th^ | 2.93 |  |  | 0.00 (-0.23, 0.23) | 0.98 | -0.15 (-0.34, 0.04) | 0.13 |
| 50^th^ | 4.32 |  |  | 0.11 (-0.25, 0.46) | 0.55 | 0.13 (-0.27, 0.53) | 0.54 |
| 75^th^ | 7.99 |  |  | 0.93 (0.13, 1.74) | 0.02 | 1.86 (0.72, 3.00) | 0.001 |
| 90^th^ | 15.56 |  |  | 3.25 (0.74, 5.75) | 0.01 | 8.21 (4.65, 11.78) | <0.001 |
| **Moderate GP Service Group (7-11 GP services)** | | | | | | | |
| 10^th^ | 3.15 | 0.01 (-0.38, 0.41) | 0.95 | -0.26 (-0.75, 0.23) | 0.30 | -0.19 (-0.61, 0.24) | 0.38 |
| 25^th^ | 4.15 | 0.09 (-0.66, 0.84) | 0.82 | 0.01 (-0.58, 0.59) | 0.98 | 0.18 (-0.40, 0.75) | 0.55 |
| 50^th^ | 6.82 | 1.17 (0.25, 2.10) | 0.01 | 0.57 (-0.26, 1.41) | 0.18 | 0.65 (-0.03, 1.33) | 0.06 |
| 75^th^ | 11.35 | 1.14 (-0.62, 2.89) | 0.20 | 1.55 (0.06, 3.04) | 0.04 | 3.19 (1.60, 4.79) | <0.001 |
| 90^th^ | 20.62 | 1.47 (-2.48, 5.41) | 0.47 | 5.29 (0.99, 9.60) | 0.02 | 9.12 (3.85, 14.40) | 0.001 |
| **High GP Service Group (12-22 GP services)** | | | | | | | |
| 10^th^ | 5.65 | 0.56 (-0.77, 1.90) | 0.41 | -0.38 (-1.41, 0.65) | 0.47 | -0.26 (-1.36, 0.83) | 0.64 |
| 25^th^ | 9.01 | -0.10 (-1.74, 1.55) | 0.91 | 0.05 (-1.42, 1.53) | 0.94 | 0.06 (-1.37, 1.49) | 0.94 |
| 50^th^ | 13.27 | 1.85 (-0.30, 4.00) | 0.09 | 2.08 (0.25, 3.91) | 0.03 | 1.30 (-0.65, 3.25) | 0.19 |
| 75^th^ | 20.46 | 1.76 (-2.01, 5.54) | 0.36 | 3.86 (-0.03, 7.75) | 0.05 | 2.58 (-0.95, 6.10) | 0.15 |
| 90^th^ | 41.46 | 3.33 (-2.73, 9.38) | 0.28 | 7.79 (1.20, 14.39) | 0.02 | 2.59 (-3.46, 8.64) | 0.40 |
| **Very High GP Service Group (23+ GP services)** | | | | | | | |
| 10^th^ | 9.70 | 7.95 (3.58, 12.32) | <0.001 | 4.46 (-0.36, 9.28) | 0.07 | 0.77 (-3.80, 5.35) | 0.74 |
| 25^th^ | 16.09 | 12.95 (7.86, 18.03) | <0.001 | 9.01 (2.04, 15.97) | 0.01 | 4.99 (-0.90, 10.88) | 0.10 |
| 50^th^ | 43.30 | 8.42 (2.92, 13.91) | 0.003 | 5.88 (-0.55, 12.31) | 0.07 | 3.10 (-3.86, 10.06) | 0.38 |

Note: # Weeks lost at this percentile calculated at the reference value of categorical variables and minimum number of GP services. Note: ^ Coefficient refers to difference in weeks lost compared to those with complete continuity of care (UPC = 1)	Note: * A score of 0.75-0.99 is not possible on the Bice-Boxerman metric for claims with 4-6 services	COCI: Bice-Boxerman Continuity of Care Index, CI: Confidence Interval, GP: General Practitioner
